# Upscaling health worker training on sepsis in South Eastern Nigeria using innovative digital strategies: an interventional study

**DOI:** 10.1101/2022.06.22.22276742

**Authors:** Akaninyene Otu, Obiageli Onwusaka, Daniel E. Otokpa, Ukam Edadi, Ubong Udoh, Peter Yougha, Chinelo Oduche, Okey Okuzu, Shevin T Jacob, Jamie Rylance, Emmanuel Effa

**Author notes:** **Corresponding Author:** E mail (EE). These authors contributed equally to this work (Akaninyene Otu, Emmanuel Effa).

## Abstract

**Introduction:** Sepsis is a leading cause of morbidity and mortality worldwide. In low- to middle-income countries (LMICs) such as Nigeria, the disproportionately high mortality rate is linked to lack of awareness, poor recognition, and late implementation of sepsis care bundles among health workers. Training of health workers using digital platforms may improve knowledge and skills and lead to better patient outcomes.

**Methods:** This Cross-sectional study involved developing and deploying a sepsis module through an innovative application (FHIND/ARCS Sepsis tutorial app) to doctors in Calabar, Nigeria. We assessed quantitative pre- and post-intervention knowledge scores for those completing the electronic training module on sepsis between both assessments. A user satisfaction survey evaluated the content of the tutorial and the usability of the app.

**Results:** One hundred and two doctors completed the course. There were more males than females (58.8% versus 41.2%). Over half (52%) were junior doctors, a minority were general practitioners and house officers (3% and 5%, respectively) and 72.6% had practiced for periods ranging from one to 15 years post qualification. Gender and age appeared to have no statistically significant association with pre- and post-test scores. The oldest age group (61-70) had the lowest mean pre- and post-test scores while general practitioners had higher mean pre- and post-test scores than other cadres.

The majority (95%) of participants recorded higher post-test than pre-test scores demonstrated by a statistically significant overall increase in mean scores (25.5% ±14.7, P<0.0001).

Participants were satisfied with the content and multimodal delivery of the material and found the app useable.

**Conclusion:** Digital training in sub-Saharan Africa is feasible and can sustainably close the critical knowledge gap required to respond more effectively to medical emergencies such as sepsis in LMIC settings.

## Introduction

Sepsis, a syndrome of life-threatening organ dysfunction due to a dysregulated host response to infection [1], is a leading cause of morbidity and mortality worldwide and particularly in Africa where awareness is low, and resources are limited [2]. Sepsis-related deaths in sub-Saharan Africa (sSA) reflect underlying political, poverty, health inequity and health system challenges across Africa [3].

There is a dearth of population incidence data for sepsis in sSA. However, data from a few available cohort studies suggest that the case fatality rates of sepsis in sSA are higher than in high-income settings [4,5]. The Global Burden of Disease 2020 study estimates 16.7 million cases of sepsis in sSA annually, accounting for up to 45% of all deaths in many those countries [6,7].

Reduction in sepsis-related mortality linked to the implementation of the sepsis-6 care bundle has been a major priority worldwide [8]. Sepsis-6 is a clinical care bundle comprised of six processes namely: blood culture, full blood count and lactate estimation, intravenous (IV) fluid challenge, IV antibiotic(s) administration, urine output monitoring and oxygen therapy. When completed within the first one hour of admission, these six processes have been associated with improved mortality.^9^ Every hour of delay in antibiotic administration has been associated with an 8% increase in mortality among patients with septic shock [10]. A recent systematic review with potential selection and information bias and confounding suggests that the evidence for the recommended timings is not robust [11].

Nonetheless, these recommendations reflect the need for heightened awareness and urgency in the initial evaluation and target-driven approach to the management of sepsis [12]. Lack of awareness of early symptoms and their seriousness has been linked to delayed health seeking behaviour among people in several African contexts [13,14,15]. In high-income countries, public awareness of sepsis has been increasing due to concerted efforts by several groups such as the Surviving Sepsis Campaign and a multitude of national regulators and expert bodies [14,15]. However, in low- and middle-income country (LMIC) settings such as Nigeria, a low level of awareness of sepsis may be contributing to poorer outcomes.

Medical personnel can introduce further delays due to lack of knowledge of sepsis. Although the recommendations for sepsis recognition and management have evolved over the years, it is unclear if healthcare workers especially doctors are fully aware of these changes. Presently, there are no local guidelines for management of patients with sepsis and critically unwell patients in Cross River State (CRS) located in south-eastern Nigeria. Consequently, the medical care of these groups of patients may not reflect evidence-based international standards.

In this study, we designed and implemented a digital health educational module on sepsis (FHIND/ARCS Sepsis tutorial app) aimed at improving the knowledge of frontline medical doctors across CRS on the diagnosis and management of patients presenting with sepsis. This was complemented by the deployment of education, information, and communication (EIC) materials for the general population to raise awareness on and reduce the likelihood of death from sepsis in CRS.

## Materials and methods

### Study setting

CRS is in Southeast Nigeria and has a population of approximately 3,866,300 people who are served by three tertiary level facilities, 16 secondary level facilities (general hospitals) and 909 primary level facilities. Approximately 70% of clients in CRS access care from public facilities. The University of Calabar Teaching Hospital (UCTH) is a 500-bed hospital that has two intensive care units (adults and neonatal) and serves as the main specialist referral hospital in CRS. Between 2012 and 2017, nearly 50,000 patients were admitted. Of about 2200 deaths, 26.0% were attributable to sepsis [18]. Sepsis has also been reported to cause high maternal mortality at the UCTH. [19,20].

### Conceptual framework

The theoretical underpinning for this study is linked to the three-layered behaviour change wheel which comprises opportunity, capability, and motivation [21]. These factors are in turn driven by attributes such as physical, physiological, psychological, and social behaviours. In addition, the delivery of interventions such as training, education and incentivization of practice enable behavioural change which ultimately leads to outcomes such as adoption of guidelines and better service provision [22]. In the context of a healthcare setting, deployment of a digital training platform against the backdrop of current suboptimal behaviour in the care of sepsis patients at the UCTH should lead to a change in better infection prevention and control (IPC) practices, earlier diagnosis and quick responses, as well as appropriate antibiotic prescribing for sepsis patients. Ultimately, these improvements should lead to a reduction in morbidity and mortality (Fig 1).

**Fig 1.**
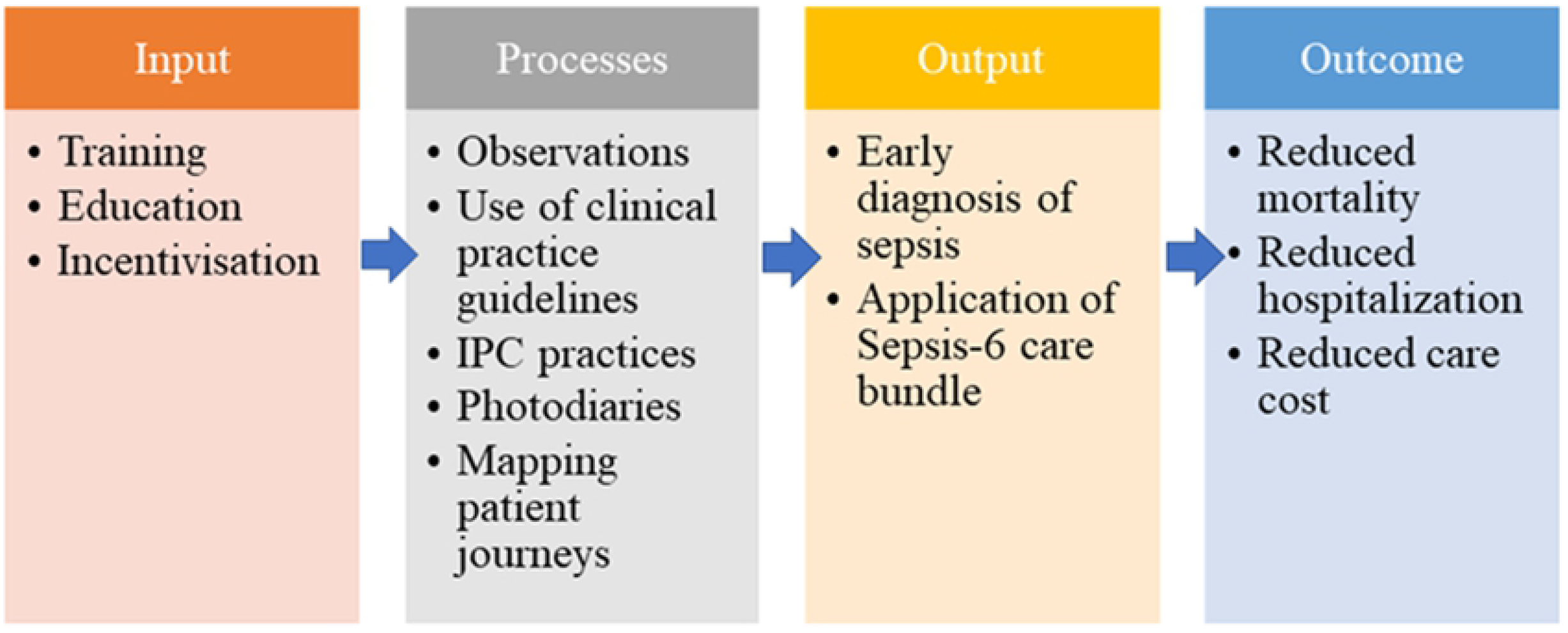
Behaviour change wheel framework: Incorporates sources of behaviour, intervention targets and policy outcomes

### Design

Our group the Foundation for Healthcare Innovation and Development (FHIND) leveraged its extensive experience in developing and deploying low-cost and effective digital training materials for frontline health workers to address diseases such as Ebola Virus Disease and COVID-19 [23,24]. This study involved quantitative pre- and post-intervention knowledge assessments for the health workers who had to complete the electronic training module on sepsis between both assessments. The assessment was followed by a user satisfaction survey after which the health workers were eligible for Continuing Professional Development (CPD) credit points issued by the Nigerian Medical Association (NMA), CRS branch. The course contents had been assessed and accredited by the NMA, CRS. The representative target sample size needed, to achieve the study objectives and sufficient statistical power, was calculated with a sample size calculator. The sample size calculator arrived at 100 participants, using a margin of error of ±9.2%, a confidence level of 95%, a 50% response distribution, and 777 people.

### Implementation

The FHIND/ARCS Sepsis tutorial app was made freely available to the 777 doctors registered on the databases of the CRS chapters of the NMA, Medical and Dental Consultants’ Association of Nigeria (MDCAN), Association of Resident Doctors (ARD), and Association of General and Private Medical Practitioners of Nigeria (AGPMPN) between 24 October and 20 December 2021. All doctors who were currently practicing in CRS were eligible to participate in the study. The tutorial app (Fig 2) was designed using the video training (VTR) mobile training platform by InStrat Global Health Solutions (https://instratghs.com/), an indigenous information technology company. The tutorial app was based on content developed by the Foundation for Healthcare Innovation and Development (FHIND) (www.fhind.org). It consisted of a narrated slide set and two complementary videos lasting five minutes each on sepsis. Upon reading study information (Supplementary File), interested participants completed a consent form electronically. User logins were then created and sent to the participants via WhatsApp, email and SMS by the staff of InStrat to download onto their android mobile phones and computers. The participants then used the learning material at their convenience, and their progress was tracked automatically. We estimated that it would take the participants at least 90 minutes to complete the module. InStrat and FHIND staff were involved in monitoring of responses, pre-test, training module completion and post-test assessments, and support was provided to the participants via WhatsApp and telephone. An end-of-course evaluation survey was sent out to all participants who completed the sepsis training course using Google forms. This survey was aimed at understanding how effective the FHIND/ARCS Sepsis tutorial app was for disseminating vital information to the health workers. Participants were eligible for certificate of completion and/or CPD certificates upon scoring at least 80% in the post-test assessment and completing the course evaluation survey.

**Fig 2.**
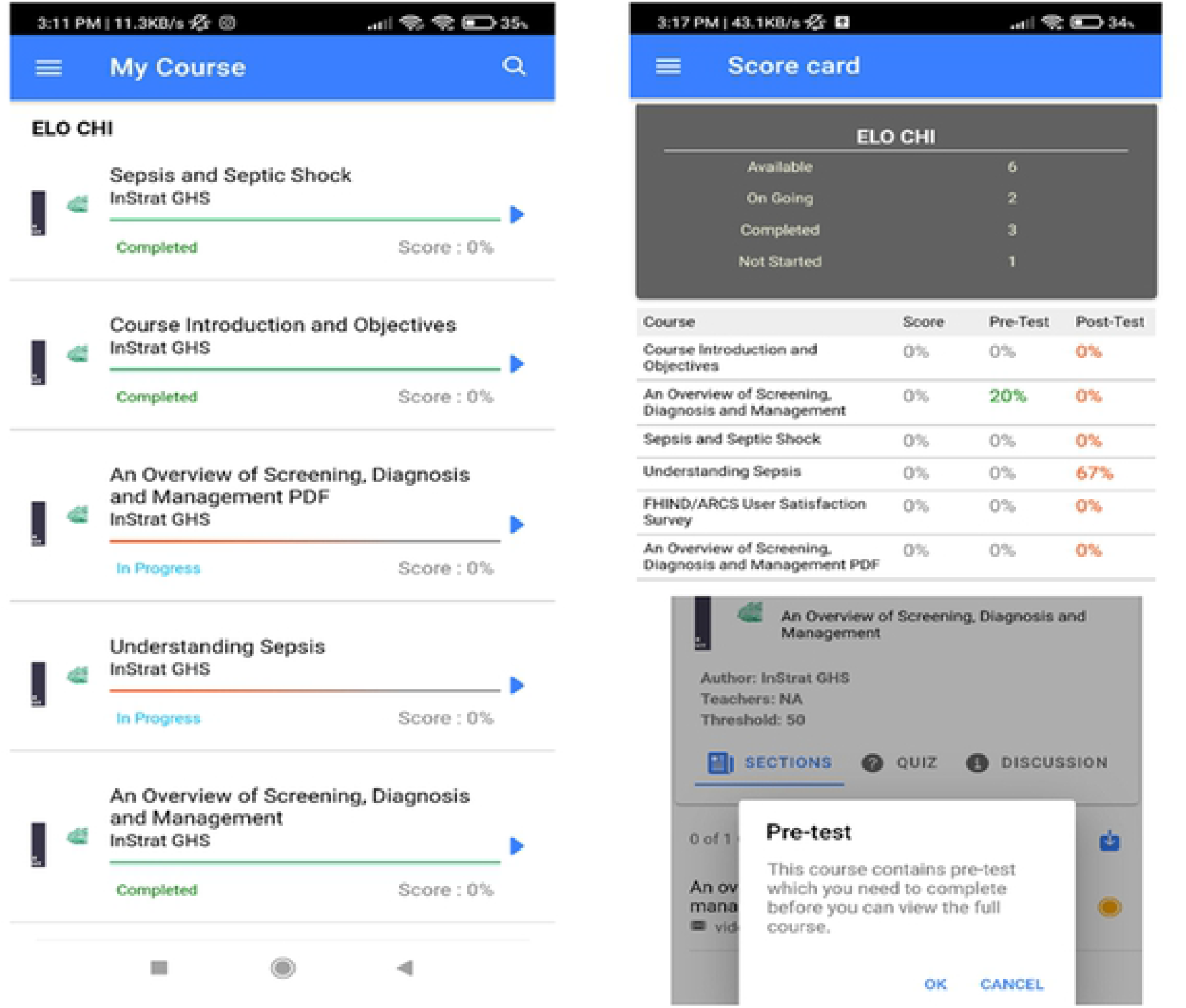
FHIND/ARCS Sepsis tutorial app

## Statistical analysis

The socio-demographic characteristics of the participants were described using relative frequency. To test for homogeneity of variances and normality of distribution of the pre-test and post-test scores, we utilised the Levene’s test and Shapiro-Wilk test, respectively. Logarithmic transformation was performed on the pre- and post-test scores to allow use of parametric tests for analysis. Paired Student’s t-test was used for comparing mean pre- and post-test scores of the participants while analysis of variance (ANOVA) was used to compare them between groups. A cut-off of P < 0.05 was used to determine significance. Only participants who completed both the pre and post-tests were included in the analyses.

Analyses were conducted using SPSS Statistics for Windows, Version 20.0. Armonk, NY: IBM Corp. IBM Corp.

## Ethics approval

The study protocol was approved by the Health Research and Ethics Committee of the UCTH with assigned number UCTH/HREC/33/579.

## Results

A total of 777 requests for participation were sent out via email to medical doctors across several affiliates of the NMA and facilities in CRS. Of these, 110 consented to participate in the study out of which 102 completed the study. There were more males than females (58.8% versus 41.2%), over half (52%) were junior doctors, a minority were general practitioners and house officers (3% and 5%, respectively) and 72.6% had practiced for periods ranging from one to 15 years post qualification (Table 1).

**Table 1.** Characteristics of study respondents (Medical Doctors: n=102)

| Characteristics | | Frequency<br>n=102 | Percent<br>(%) | Pre-test score<br>(Mean ± S.D.) | Post -test score<br>(Mean ± S.D.) |
| --- | --- | --- | --- | --- | --- |
| Gender | Male | 60 | 58.8 | 59.5 (14.2) | 83.3 (6.0) |
|  | Female | 42 | 41.2 | 56.7 (11.5) | 84.5 (5.3) |
| Age | 21-30 | 11 | 10.8 | 61.5 (15.6) | 81.2 (3.9) |
|  | 31-40 | 48 | 47.1 | 58.7 (14.0) | 84.7 (6.1) |
|  | 41-50 | 31 | 30.4 | 56.6 (11.8) | 82.7 (4.7) |
|  | 51-60 | 9 | 8.8 | 59.1 (12.3) | 87.3 (7.7) |
|  | 61-70 | 3 | 2.9 | 55.0 (10.1) | 80.0 (0.0) |
| Professional level | Consultant | 27 | 26.5 | 56.9 (10.8) | 82.9 (4.5) |
|  | General practitioner | 3 | 2.9 | 71.0 (10.1) | 97.7 (4.0) |
|  | House officer | 5 | 4.9 | 61.4 (14.5) | 80.0 (0.0) |
|  | Medical officer | 14 | 13.2 | 52.1 (8.7) | 85.6 (7.2) |
|  | Resident doctor | 53 | 52.0 | 59.7 (14.7) | 83.1 (5.1) |
| Years of practice<br>(post qualification) | 1-5 years | 15 | 14.7 | 58.9 (14.5) | 83.0 (4.8) |
|  | 6-10 years | 33 | 32.4 | 59.5 (14.9) | 85.7 (6.6) |
|  | 11-15 years | 26 | 25.5 | 60.0 (12.4) | 81.5 (3.8) |
|  | 16- 20 years | 12 | 11.8 | 44.8 (10.6) | 83.2(4.4) |
|  | 21-25 years | 8 | 7.8 | 60.6 (11.4) | 87.4 (8.2) |
|  | >25 years | 8 | 7.8 | 54.8 (9.3) | 82.5 (4.9) |

The mean pre-test score for all participants was $58.3 \pm 13.2$ , while the mean post- test score was $83.8 \pm 5.7$ . Age-wise and gender-wise, there were no statistically significant differences in pre- and post-test scores of the participants. However, there was a statistically significant difference ( $P < 0.0001$ ) in post- test scores across the different professional levels when compared with general practitioners (accounting for the highest post-test scores) and house officers (accounting for the lowest post-test scores). Furthermore, post-test scores also significantly differed ( $P = 0.037$ ) according to the number of years of practice.

The overall mean scores showed a highly significant (p<0.0001) improvement in the post-test scores of all the participants compared to their pre-test scores with a mean difference of 25.5% ±14.7 between pre- and post-test scores.

Levene’s test showed equal variance for the post test scores across the categories of age (p=0.001), professional level (p=0.000) and years of post-qualification practice (p=0.001). The Shapiro-Wilk test showed that the distribution of both the pre- and post-test scores departed significantly from normality (W =0.962, *P* < 0.05 for pre-test scores and W =0.690, *P* < 0.05 for post-test scores). Based on this result, log transformation of the data was performed to enable conducting of parametric tests.

A variable representing the difference between pre- and post-test scores was computed for all respondents. The mean of this variable was also computed for all the groups of participants and ANOVA was used to compare these means across the various groups (Fig 3). The mean difference of scores of the house officers was lower than the average of all participants while that of the general practitioners was higher than the average.

**Fig 3.**
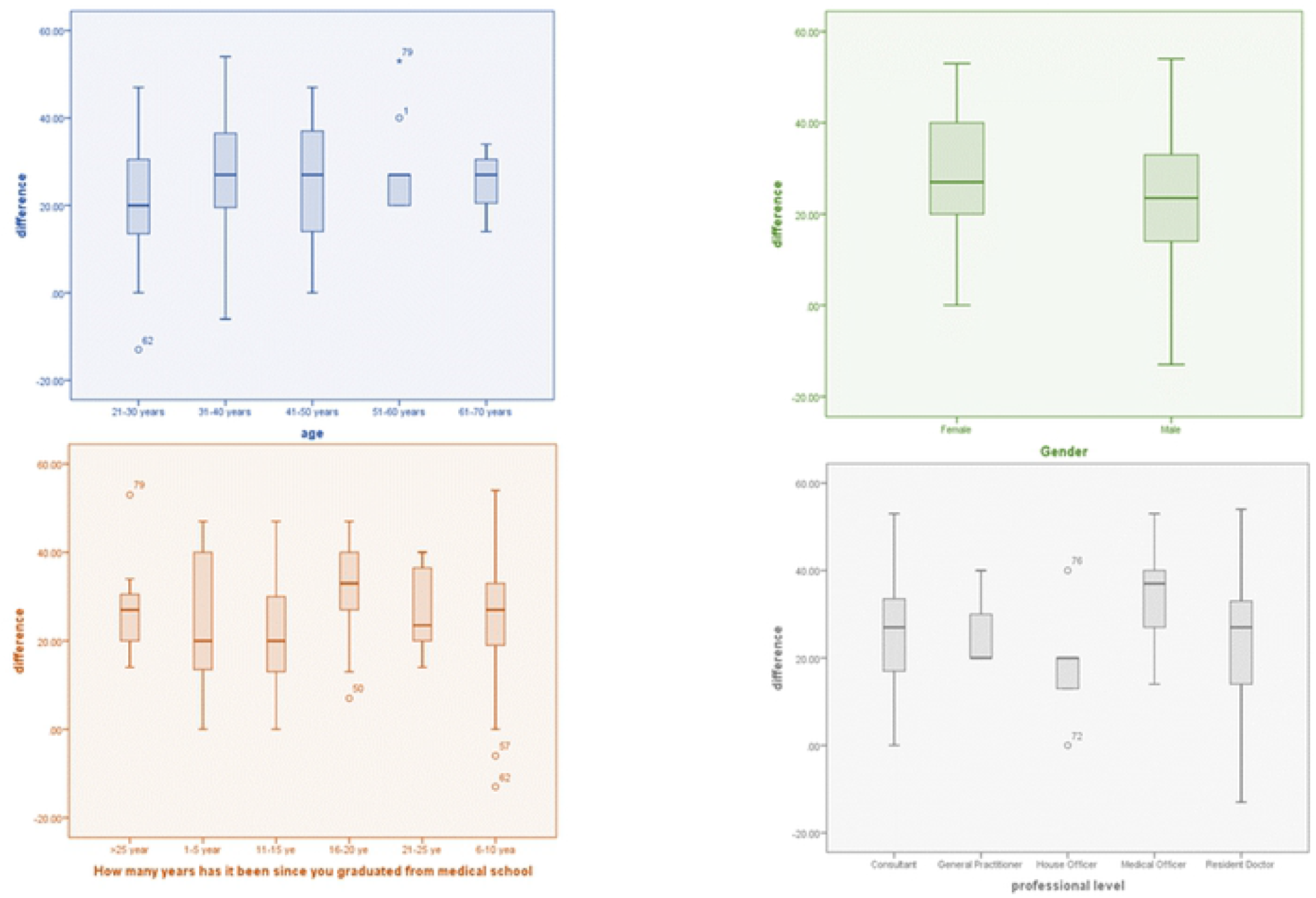
Variation in the mean differences between pre- and post-test scores

## User survey

A total of 102 participants completed the user evaluation survey [S1 Survey]. They provided responses on usability, user experience, and applicability of the app, and the knowledge derived from it. Seventy (69%) of them stated they had received some training on sepsis prior to embarking on the FHIND/ARCS course. Most respondents (73/102, 71.2%) answered that they ‘agree strongly’ that the app had provided them with a better understanding of sepsis, and 69 (67.6%) stated that the knowledge and skills obtained would be applied in the work environment. Positive feedback on usability of the app and user experience was obtained from 82 and 88 doctors, respectively. Sixty-nine of them stated that they would prefer e-learning courses to face-to-face training. A summary of the user evaluation survey is a shown in the Fig 4.

**Fig 4.**
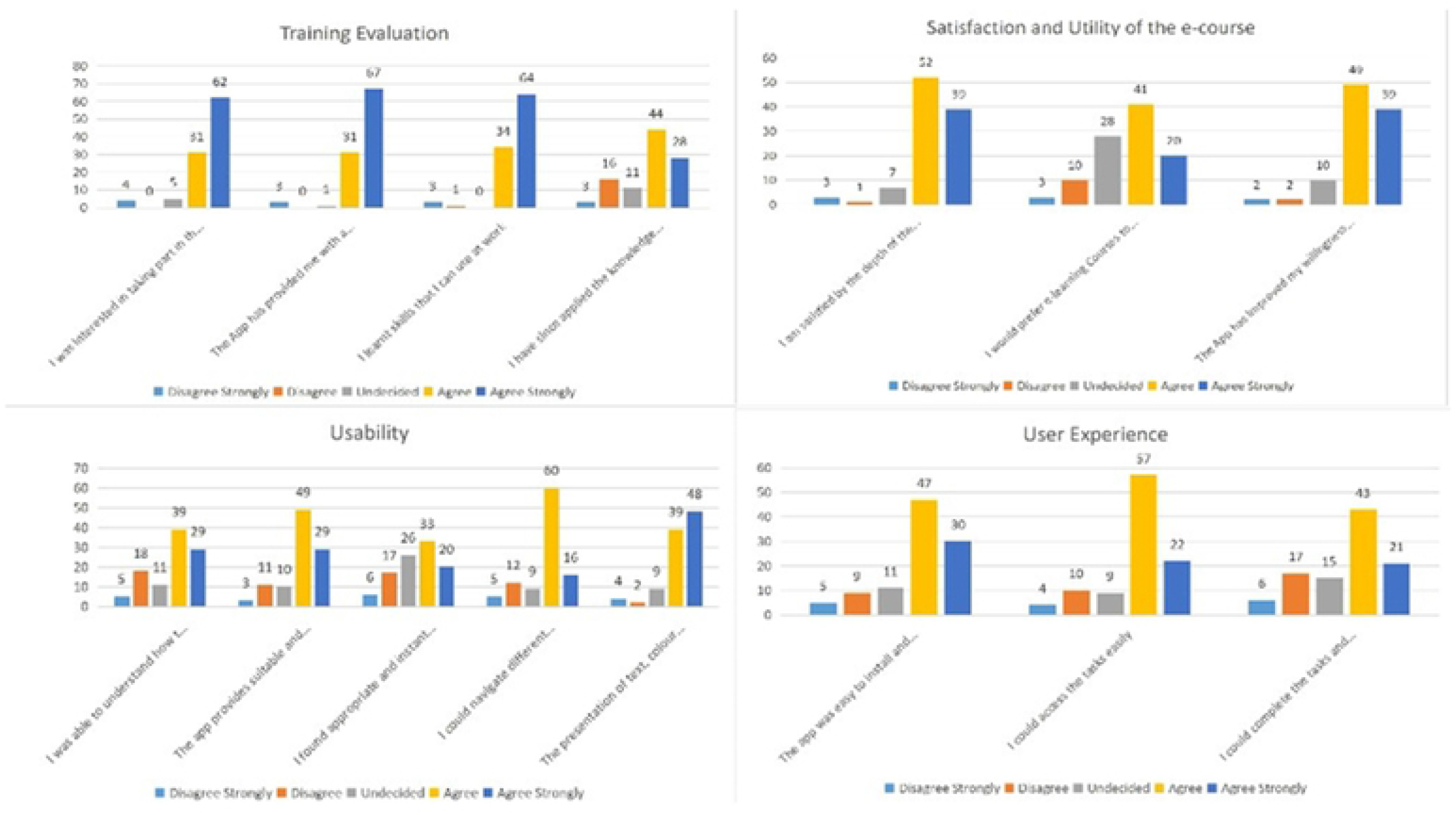
User evaluation survey.

Some illustrative feedback is provided in the box below

*Comments*

- “I enjoyed the session, the inclusion of different delivery methods - explanatory, audiovisuals and diagrams, aid in imprinting the knowledge.” (*Resident Doctor*)
- “This virtual training is an exceptional innovation to be promoted. Overall, my experience was top-notch.” (*Medical Officer*)
- “I wish to thank the team for this innovation. This is the first e-learning course made in a resource limited setting like ours, I have partaken in.” (*Resident Doctor*)
- “The course is timely and has enriched my knowledge on management of patients with sepsis/septic shock” (*Medical Officer*)

*Suggested areas for improvement*

- “The video section should be like a live demonstration not just the audio. More videos. Occasional visuals of presenters may be helpful” (*Medical Officer*)
- “Text copies of the video and audio material as not all persons are complete audio-visual learners.” (*Resident Doctor*)
- “If the modules could be downloaded and stored and the training done offline before uploading it could help those with erratic internet connectivity.” (*Consultant*)
- “Navigation through the app can be improved on especially when going back to the course page; it does not take someone back when one finishes a material. Fantastic job!” (*Resident doctor*)
- “Would follow-up short reminders and an assessment of its impact be done months after?”(*Resident Doctor*)
- “What about non-clinicians and clinicians as well, can there be more emphasis on integrated preventive strategies?” (*Resident Doctor*)

## Discussion

Our study involved the use of digital health platform to train 102 frontline doctors in CRS on sepsis with statistically significant improvements in their pre- and post-test scores. The majority of those trained reported an improvement in their understanding of sepsis and acquisition of skills relevant to their work. Many also provided positive feedback on usability of the FHIND/ARCS Sepsis tutorial app.

In Nigeria, conventional face-to-face residential training is the typical approach for providing CPD for doctors. However, this approach is fraught with financial and logistic challenges, including the need for resource persons to conduct the training, the need for travel of participants across long distances to attend the trainings and limited options for scalability [26].

The increasing availability and use of digital technologies in Nigeria offers new opportunities to support remote training. Nigeria has the fastest growing mobile market on the African continent with mobile phone subscription in Nigeria reaching 180 million in October 2019 and smart phone usage in Nigeria currently estimated at 40 million [27]. This increasing penetration provides a springboard for launching digital training for health workers on key topics such as sepsis as many frontline health workers in Nigeria own smartphones [27].

Our previous efforts utilising a similar app to train frontline health workers in Nigeria on Ebola Virus Disease [23] and COVID [24] demonstrated the feasibility and acceptability of using digital technology to deliver training programmes in low-resource settings. The findings of our study corroborate two large systematic reviews that concluded that digital training was as effective as traditional methods for improving the knowledge and skills of health workers [28,29].

Our research findings have several implications for health policy, practice, and future research. Our digital training approach holds promise for low- and middle-income country settings with minimal health budgets as it can provide high-quality, customised information to health workers in a safe, minimally disruptive, and inexpensive way. We were also able to evaluate learning and skills acquisition using the same platform which is an added advantage.

Participants appreciated the multimodal delivery strategy of the course which include a mix of texts, audio-visuals, and diagrams. Indeed, a shift from a teacher-centric to a student-centric approach has been demonstrated to motivate learners, keep them attentive and improve learning [30]. Content creation for e-learning platforms such as the one deployed here would need to take this shift into consideration for optimal learning outcomes.

Although there was significant improvement in the post-test scores, further evaluation is needed to determine whether the newly acquired knowledge and skills will translate to behaviour change in early recognition of sepsis and prompt institution of appropriate care both in the long and short term. We identified some challenges in the process of executing this intervention. First, several of the users found it challenging to download the app due to incompatibility issues. InStrat staff had to provide remote instructions to users on the WhatsApp platform to guide them through the entire process. Second, some of the users had limited storage space to run the Sepsis app on their mobile devices, and they had to be directed to delete unused/seldom used apps and old media files to create space on their devices. For subsequent deployments of this and similar training apps, it is apparent that extensive training and supportive supervision will be vital for users who are not technologically savvy. Also, power outages and poor internet connectivity [31]. are challenges to digital training that will need to be addressed before deploying such trainings at large scale.

## Conclusion

In conclusion, with improved information communication infrastructure and appropriate e-health support, digital training in sub-Saharan Africa is scalable and can sustainably close the critical knowledge gap required to respond more effectively to medical emergencies such as sepsis in LMIC settings.

## Data Availability

All relevant data are within the manuscript and its Supporting Information files.

## Author Contributions

AO, EE, ObO conceptualised the study. AO, EE, ObO and OO designed the study. AO, EE, OO, SA, ObO, DO, UE, PY, CO were involved in data collection. ObO, EE, AO, PY and CO handled the data analysis and interpretation. AO wrote the first draft of the article. EE, ObO, JR and STJ critically revised the first draft. All the co-authors reviewed and approved the version of the article to be published.

## Acknowledgements

We appreciate all the doctors who participated in this study.

